# Local genetic correlation analysis links depression with molecular and brain imaging endophenotypes

**DOI:** 10.1101/2023.03.01.23286613

**Authors:** J. Werme, E.P. Tissink, S.C. de Lange, M.P. van den Heuvel, D. Posthuma, C.A. de Leeuw

**Affiliations:** Department of Complex Trait Genetics, Center for Neurogenomics and Cognitive Research, Vrije Universiteit Amsterdam, Amsterdam Neuroscience, 1081 HV Amsterdam, The Netherlands; Department of Sleep and Cognition, Netherlands Institute for Neuroscience, an institute of the Royal Netherlands Academy of Arts and Sciences, The Netherlands; Department of Child Psychiatry, Section Complex Trait Genetics, Amsterdam Neuroscience, Vrije Universiteit Medical Center, Amsterdam UMC, Amsterdam, The Netherlands

## Abstract

Major depressive disorder (MDD) is a heritable psychiatric disorder which is considered one of the leading causes of disability world-wide. Improved understanding of its genetic component could inform novel treatment developments, but so far, gaining functional insights from genome-wide association studies has been difficult. In this study, we sought to generate hypotheses about plausible mechanisms through which genetic variants could influence MDD using a novel approach. Considering the cis-regions of protein coding genes as the loci of interest, we applied local genetic correlation analysis to study the genetic relationship between MDD and a range of brain, endocrine, and immune related endophenotypes across several modalities (tissue specific gene expression and splicing, regional brain volumes, and brain network connectivity). We identify significant genetic relations between MDD and endophenotypes within the cis-regions of multiple genes, and perform endophenotype specific enrichment analyses of the top associated genes. Our results offer potential mechanisms through which MDD related variants in these genomic regions could act, and convergent evidence from multiple endophenotypes implicate *FLOT1* as a gene which may exhibit wide-ranging pleiotropic effects and be particularly interesting for functional follow-up. Here, we have illustrated how local genetic correlation analysis applied to lower level endophenotypes has the power to prioritise genes and functional paths which warrant further investigation for their possible role in MDD aetiology.

## INTRODUCTION

Major depressive disorder (MDD), or depression, is a common but debilitating psychiatric disorder characterised by symptoms such as persistent negative mood, anhedonia, lack of motivation, and sometimes suicidal ideation^1^. With an estimated lifetime prevalence of around 11%^2^, it is considered to be one of the leading causes of disability worldwide^3^, yet so far, first line treatment options for depression are of limited efficacy^4,5^.

Depression is influenced by a combination of genetic and environmental factors^6^, with the estimated heritability obtained from twin studies ranging from 30%-50%^6^ and the additive effects of common SNPs accounting for around ∼3% of the variance in depression status^7^. The advent of large scale, unbiased investigation of the genetic component of depression in the form of genome-wide association studies (GWAS) was expected to provide substantial insights into the biological mechanisms that underlies it, yet discerning functionally relevant information and prioritising causal genes has proven difficult due to the highly polygenic nature of the disorder^6,8^.

To date, the largest GWAS meta-analysis of depression identified 178 genomic risk loci and biological processes related to for example nervous system development and neuronal signalling^9^. While the findings from gene-set enrichment analysis can provide additional biological information related to SNP and gene level results as a whole, such results are largely non-contextual, and what the association between depression and variants in a particular gene means for its aetiology is not always clear. One way of gaining functionally relevant information from GWAS, while also prioritising genes which may be likely causal, is to examine pleiotropy between depression and relevant endophenotypes. Integration of expression quantitative trait locus (eQTL) data is a relatively common approach^10–13^, but the pleiotropy between depression and regional brain volumes, or brain networks, has not yet been systematically examined on the gene level, nor across brain imaging and molecular endophenotypes simultaneously. Though doing so could provide important clues about the underlying biological mechanisms that govern depression.

Here, we rely on the currently largest publicly available GWAS summary statistics from Howard et al^7^, which represents a combination of broader self-report and clinically derived depression phenotypes. With this, we employ a novel strategy to extract functional information from the depression GWAS signal and prioritise relevant genes and endophenotypes via local genetic correlation. Using LAVA (Local Analysis of [co]Variant Association)^14^, we define our genomic loci as the cis-regions of all protein coding genes and, within these genes, evaluate the local genetic relationship between depression and endophenotypes across three modalities: tissue specific mRNA expression and splicing from 18 tissues, the volumes of 56 cortical and subcortical brain regions, as well as the structural and functional connectivity within 7 brain networks (see **Fig. 1** for an overview). We look specifically for genes where we detect significant genetic correlations across multiple levels of function, as these genes might be particularly interesting for functional follow-up due to potential wide-ranging effects on the system. We also perform endophenotype specific enrichment analyses of the top 1% of analysed genes for all endophenotypes separately with the aim of providing additional biological context to any detected gene-set associations.

**Figure 1.**
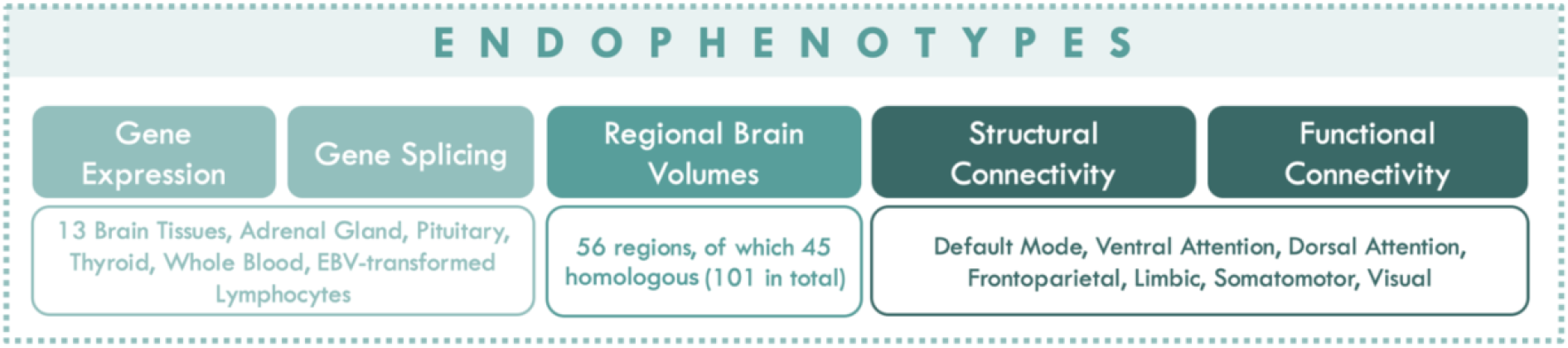
Schematic overview of analysed endophenotypes. Considering the cis-regions of all protein coding genes as our genomic loci of interest, we sought to elucidate the function of the depression GWAS signal by detecting genic regions within which there is likely pleiotropy between depression and endophenotypes across three modalities: the expression and splicing of mRNA transcripts across brain, endocrine, and immune tissues, regional brain volumes, and functional and structural brain network connectivity.

Our results show significant genetic correlations between MDD and endophenotypes across all analysed levels of function. The genetic correlations between depression and the tissue specific splicing and expression tended to be concentrated within the major histocompatibility complex (MHC) and involved almost all analysed tissues. From the regional brain volume analyses, we detected significant genetic correlations with brain regions primarily within and around the basal ganglia, but also some frontotemporal regions and cerebellar vermal lobules. Significant gene set associations were also based on the top genes from the analyses with the precentral gyrus, palladium, putamen, and the pars triangularis, which were related to cellular response to metal ions, growth regulation, triglyceride lipase activity, and hyaluronan metabolism. Finally, genetic correlations between depression and the structural connectivity within the visual network were also detected, and convergent evidence from multiple endophenotypes highlighted *FLOT1* as a gene of particular interest due to the large number of associations detected in this region.

## RESULTS

### Analysis procedure and overview

For all endophenotypes, we used LAVA^14^ to conduct local genetic correlation analyses with depression, testing the shared genetic signal at individual genomic loci. Locus boundaries were defined using the genomic coordinates of all protein coding genes (*N* = 18,380), adding 1 megabase windows around the gene transcription start sites to encompass the entire cis-region (note that we chose to focus only on the cis-regions as opposed to the entire genome due to the lack of publicly available genome-wide eQTL data from GTEx).

To filter out non-associated loci which are not of interest for the genetic correlation analysis, we applied the univariate test in LAVA to evaluate the local heritability for all phenotypes and genes. Across all endophenotypes analysed in this study, we performed a total of 72,701 local bivariate genetic correlations analyses, resulting in a corrected statistical significance threshold of 6.88e-7 (.05 / 72,701). Following the bivariate local genetic correlation analyses, we performed stratified gene-set enrichment analyses for all the individual endophenotypes separately, selecting the top 1% of associated genes and evaluating whether these were over-represented within 7,246 MSigDB^15^ gene sets. This stratification allows the results to be interpretable in relation to the specific endophenotype, providing additional context to any detected gene-set associations.

### Molecular endophenotype analyses suggest MHC signal affects expression and splicing

To determine which of the GWAS depression signal could be accounted for by effects on gene expression and splicing in relevant tissues, we used data from the GTEx consortium (v8)^16^ to model the local genetic relationships with depression across 18 tissues related to the brain, endocrine, and immune systems (**Fig. 1**). We pre-selected tissues that we judged to be of greatest relevance to depression aetiology based on the existing literature, which has focused primarily on the brain^17^, but also implicated the endocrine (adrenal gland, pituitary, thyroid) and immune systems (whole blood, EBV cells)^18^.

Correcting for the number of bivariate local genetic correlation tests performed across all genes and endophenotypes in this study (*p* < 6.88e-7), we find a total of 19 significant associations with the expression of 18 genes across 6 tissues, as well as 36 significant associations with the splicing of 21 genes across 14 tissues (**Fig. 2a & 2b**). Significant genetic correlations with depression from both the expression and splicing endophenotypes were primarily concentrated within the MHC, which is also the location of one of the major signal peaks in the analysed depression summary statistics (see **Suppl. Fig. 1**). Although there are two other peaks in the depression GWAS data with similar signal strengths on chromosome 1 and 5, no significant genetic correlations were found with the gene expression or splicing in these regions. This could be due to for example time specific regulatory effects, lack of power, or relevant risk variants not being present in a large enough subset of the GTEx sample. We note that while the MHC is a region home to complex and long-range linkage disequilibrium (LD), the LD between SNPs within a given gene is accounted for here (see Werme et al^14^ for more detail). However, long range SNP LD from beyond the gene cis-region are not, meaning that genetic effects from outside the analysed genes can confound the associations as well. We also note that some amount of overlap exists between adjacent genes, which too can lead to correlated results.

**Figure 2.**
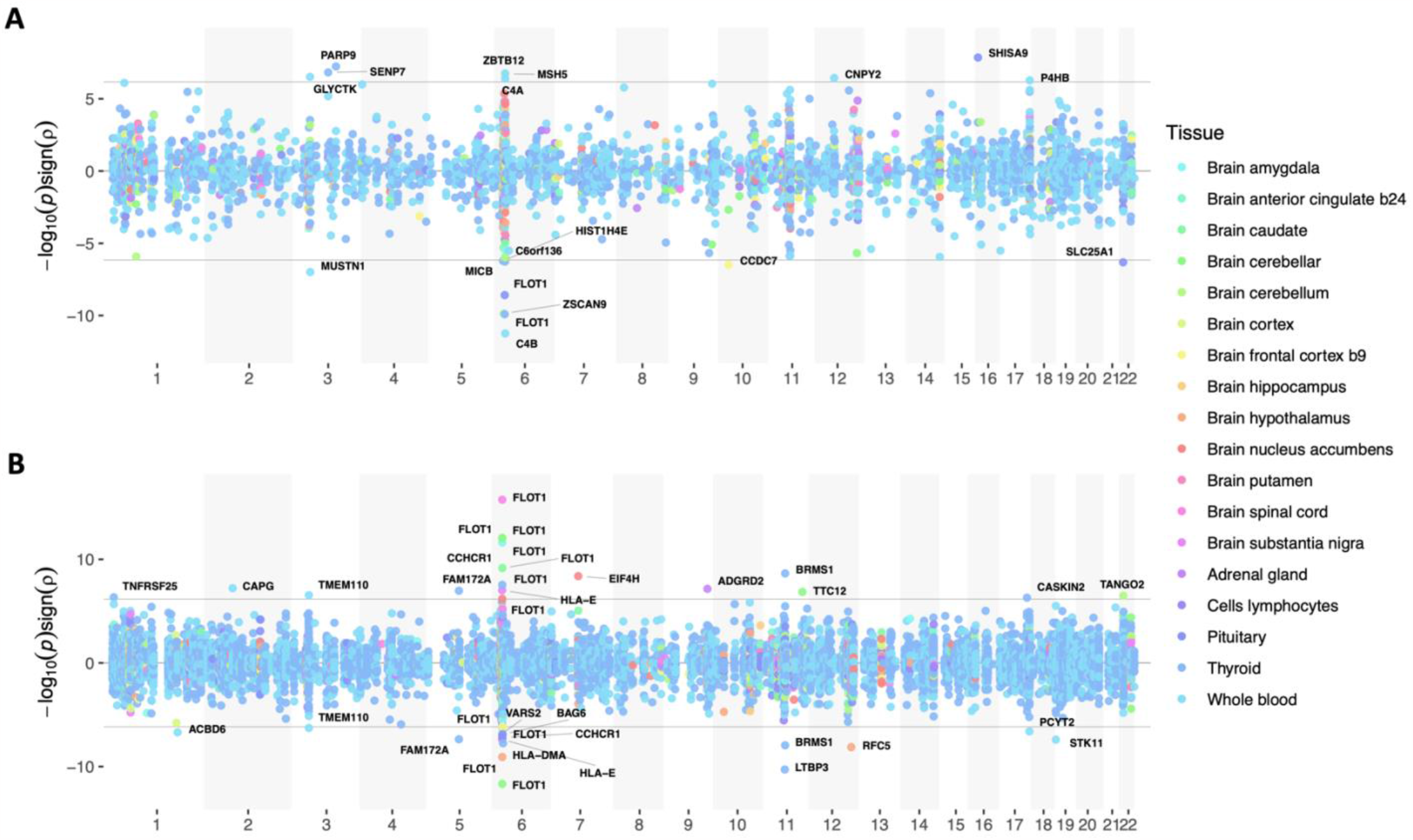
Overview of results from local genetic correlation analyses with gene expression and splicing across all 18 tissues. Panels **A** and **B** show the -log10 p-values for the local genetic correlations with gene expression and splicing respectively, scaled by the direction of the association (y-axis) and ordered according to chromosomal location (x-axis). Each point has been coloured according to tissue (see legend). Note that the individual splice variants have been omitted in panel **B** but are listed in **Suppl. Table 1** along with a complete overview of all significant results from the expression and splicing analyses.

From both the splicing and expression analyses, two of the analysed genes harboured significant genetic associations in more than one tissue: *CCHCR1* and *FLOT1. CCHCR1* showed significant associations in two tissues (brain caudate & cells lymphocytes) while *FLOT1* in as many as eleven (brain amygdala, brain caudate, brain cerebellar hemisphere, brain cerebellum, brain frontal cortex BA9, brain hypothalamus, brain nucleus accumbens, brain spinal cord, cells lymphocytes, pituitary, thyroid) (see **Suppl. Table 1** for an overview of the exact splice variants significant for all detected genes).

Of all molecular endophenotype associations, the most significant genetic correlation with MDD was found for the chr6:30,740,798-30,741,190 splice variant of *FLOT1* in the spinal cord (*r*_g_ = .74, *p* = 1.77e-16).

### Local genetic correlations with regional brain volumes implicate basal, frontotemporal, and cerebellar regions

Genome-wide association summary statistics for regional brain volumes were obtained from Zhao et al (2019)^19^. This data set contained GWAS summary statistics for a total of 56 brain regions of which 45 were hemispheric homologous regions, which were analysed separately in the original study. Given the often substantial local genetic correlations that we observed for the homologous regions, we used a meta-analysis procedure to combine the local genetic signals for these regions in loci where the explained variance for their local *r*_g_ exceeded .9, or the confidence intervals for its explained variance included 1 (see **Methods** for details).

Correcting for the total number of bivariate tests performed across all endophenotypes in this study (*p* < 6.88e-7), we found 37 significant local genetic correlations with the volumes of 14 brain regions (**Fig. 3**). Here, just over half of the brain regions with which significant genetic relationships with MDD were detected were located within or immediately adjacent to the basal ganglia (i.e. basal forebrain, brain stem, caudate, lateral ventricle, pallidum, putamen, thalamus, ventral diencephalon), an area often regarded as central to emotion processing and reinforcement learning^20^ which has been frequently implicated as a key area involved in the susceptibility to depression^21^ and other psychiatric disorders^21,22^.

**Figure 3.**
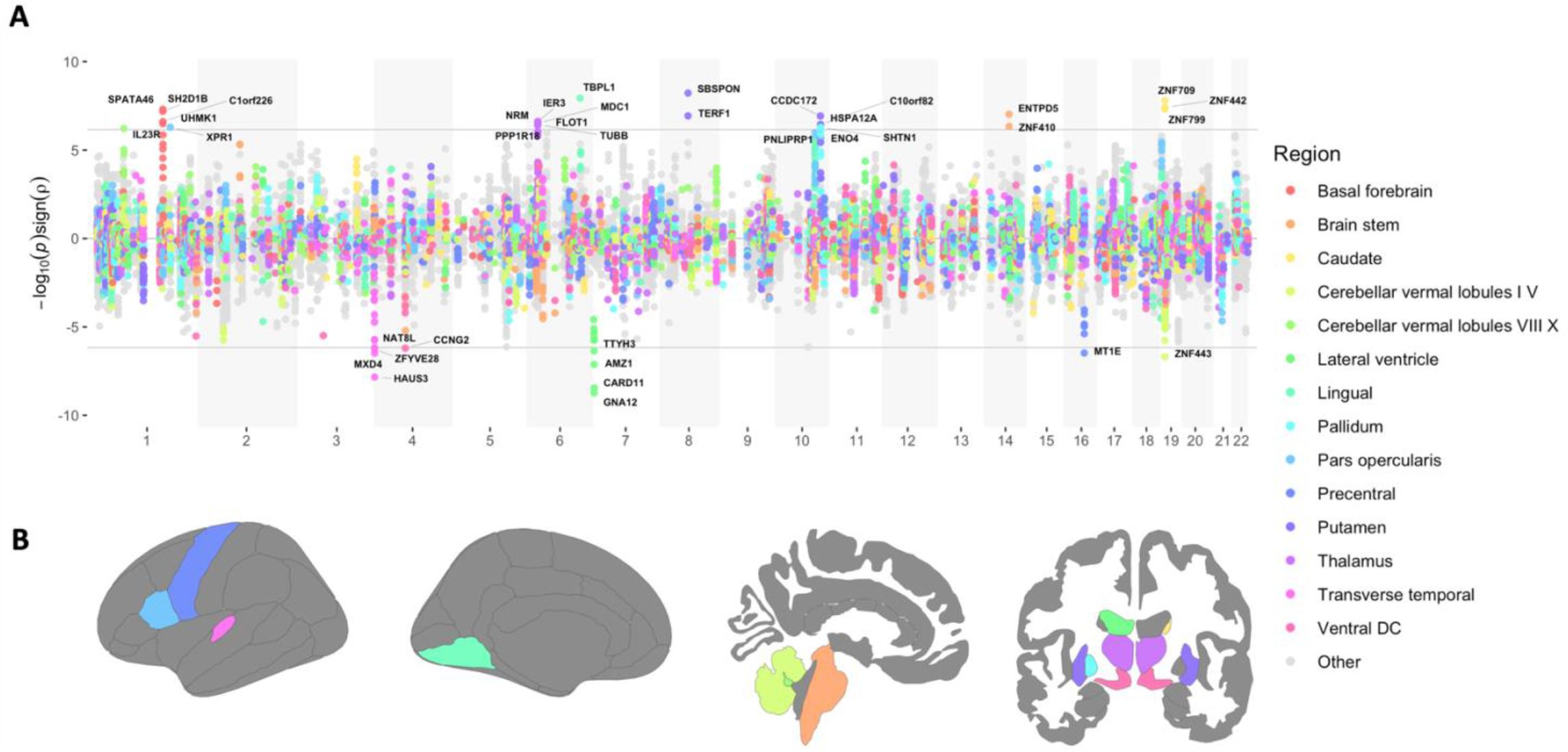
Overview of results from local genetic correlation analyses with regional brain volumes. Panel **A** shows the results from all analysed genes and regions, with points representing the -log10 p-values scaled by the direction of the genetic correlations (y-axis), which have been ordered according to chromosomal location (x-axis). Note that for the purpose of readability, only the regions within which we detected at least one significant gene have been coloured, and lateralisation of hemispheric homologous regions is not indicated (but a detailed overview of all significant hits can be found in **Suppl. Table 2**). Panel **B** shows a graphical illustration of the regional brain volumes with at least one significant hit from the local genetic correlation analyses.

Significant genetic correlations with MDD were also detected in two of the cerebellar vermal lobules [1/5 & 8/10], the lingual gyrus, the precentral gyrus, pars opercularis (part of the inferior frontal gyrus), and the transverse temporal gyrus (see **Fig. 3B** for a graphical illustration of all relevant brain regions). Though these regions have received comparatively little attention relative to the basal ganglia, they have also been implicated depression in the past^23,24^. Overall, the results presented here offer the hypothesis that the shared genetic signal within these gene cis-regions could potentially be a driving factor in the relationship between these brain regional volumes and MDD which has been shown in previous neuroimaging studies. But since the results from this study are merely observational, evaluation of this hypothesis using alternative methodologies is necessary before any such relationship can be confidently established.

Unlike the genetic correlations detected from the molecular endophenotype analyses, significant associations with the regional brain volumes tended to be more dispersed across the genome with no gene showing significant genetic correlation between depression and more than one brain region. Thus, the genes tended to be more region specific as compared to the expression and splicing analyses where the same genes tended to show genetic correlation across multiple tissues. Additionally, within the MHC, which was the region with the greatest number of significant associations from the expression and splicing analyses, only six significant correlations were found (between MDD and the left thalamus specifically). Notably, however, these included the *FLOT1* gene, which was the most frequently associated gene from the splicing and expression analyses (**Fig. 2**).

### Local genetic correlations with brain network connectivity highlight genes associated with stress hormone regulation and circadian rhythm

We used GWAS summary statistics for the connectivity within different brain networks from Tissink et al (2023)^25^ measured with resting-state functional MRI (functional) and diffusion-weighted MRI (structural) to evaluate whether the gene-level local genetic signal in depression is related to that of functional and structural brain network connectivity across seven networks (as well as the global connectivity for reference; see **Fig. 4C**).

**Figure 4.**
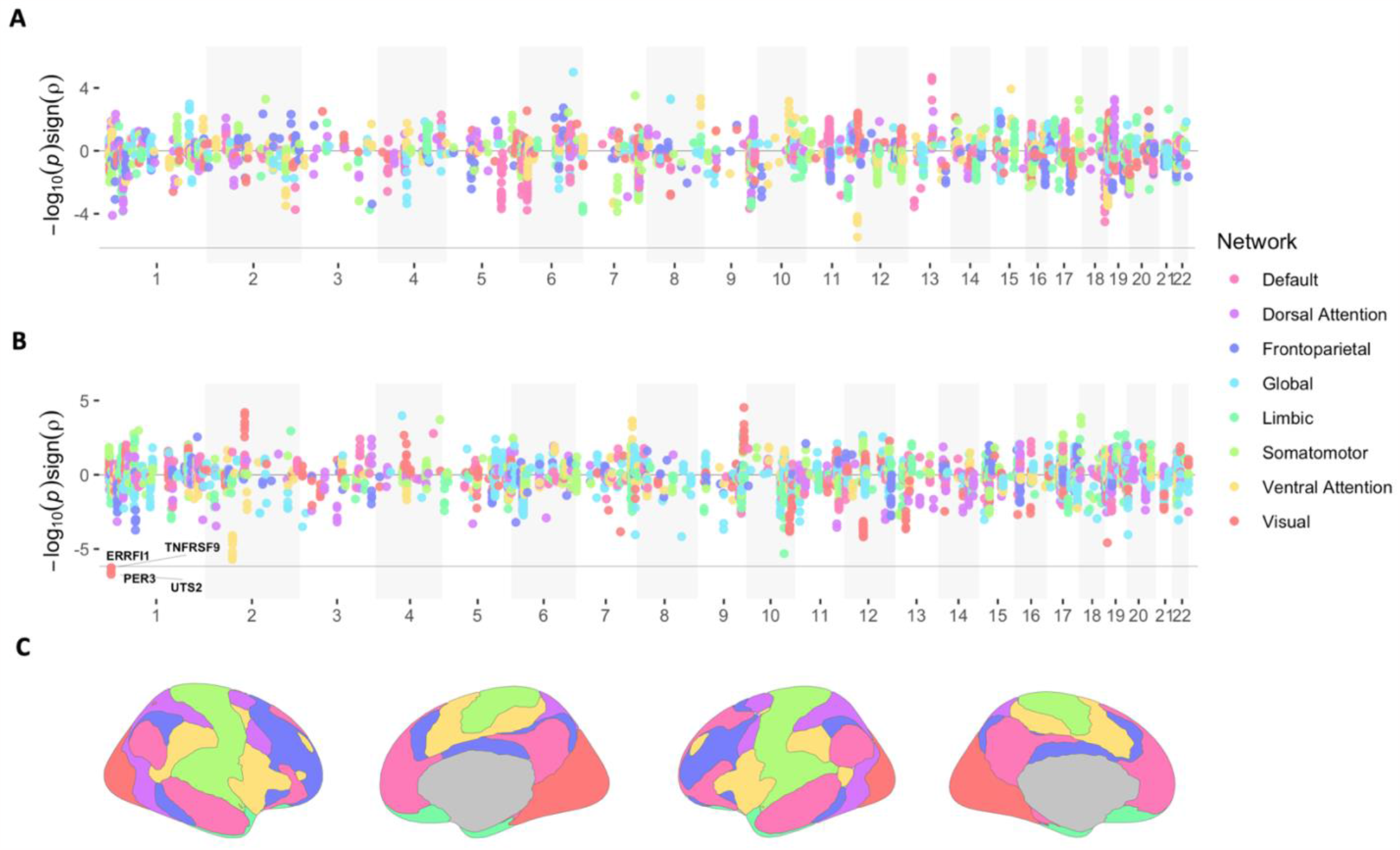
Overview of local genetic correlation analyses with functional and structural brain network connectivity. Panel **A** and **B** show the -log10 p-values from the genetic correlation analyses with the functional and structural network connectivity phenotypes respectively, scaled by the direction of effect (y-axis), and ordered according to chromosomal location (x-axis). Panel **C** shows an overview of the different brain regions related to each network, which have been differentiated by colour (see legend).

Among all analyses, we only detected significant associations with the structural connectivity within the visual network (*p* < 6.88e-7; **Fig. 4A, 4B**), where the top associated genes, *UTS2* and *PER3*, are known to be involved with functions such as stress hormone regulation and circadian rhythm^26,27,28^.

That the visual network is the only network with significant hits is not consistent with the literature, which tends to implicate networks related to higher order cognition or emotional processing^29^ in MDD, such as the default mode network^30^, ventral attention network^31^, frontoparietal network^32^, and the limbic network^32^. Though we note that power for the connectivity endophenotypes was generally low compared to other analysed endophenotypes (as indicated by the lack of genome-wide significant SNPs), and that in the original GWASs, the structural connectivity in the visual network had the greatest number of detected genome-wide significant SNPs compared to all other network phenotypes^25^. As such, the result observed here could potentially reflect the differential power between the networks, which, as highlighted in the original study, may relate to the phenotype definition (which averages less variable unimodal network connectivity compared to more variable higher-order network connectivity).

We do also emphasise that the multiple testing correction employed here is somewhat conservative as it does not account for neither the correlation between genes nor endophenotypes, and that we did observe several trend level associations with other networks as well. For example, there was an association within the *ALMS1* gene between depression and the structural connectivity in the ventral attention network which was just above the corrected significance threshold (*p* = 2.00e-6). This was also true within the *BORCS7* gene for the limbic network (*p* = 4.90e-06; structural), in the *LTBR* gene with the ventral attention network (*p* = 3.28e-6; functional), and in the *AIG1* gene with the global functional connectivity (*p* = 9.90e-6; see **Suppl. Table 3**). However, it remains to be seen whether these will be reliably detected using summary statistics form more well powered data sets.

### Stratified enrichment analyses highlight brain region specific MDD gene sets

To examine gene sets that might account for the shared genetic component between depression and analysed endophenotypes, we conducted stratified gene-set enrichment analyses for all endophenotypes separately. This was done by selecting the top 1% of genes from the bivariate local genetic correlation analyses for all endophenotypes individually, and employing a Fisher’s exact test to evaluate over-representation within 7,246 MSigDB^15^ gene sets (see **Methods** for more detail).

Correcting for the total number of gene sets tested across all endophenotypes (*p* < .05 / 28,959 = 1.73e-06), we detected 16 significant gene sets related to the top genetically correlated genes from 4 regional brain volume endophenotypes: precentral gyrus, pallidum, putamen, and pars triangularis (**Table 1**).

**Table 1.** Significant gene sets detected via enrichment analyses of top 1% genetically correlated genes, stratified by endophenotype.

| Region | Gene set | OR | P | Genes |
| --- | --- | --- | --- | --- |
| Precentral | GO_bp:go_cellular_response_to_zinc_ion | 4670.17 | 3.90E-21 | MT1E;MT1M;MT1A;MT1B;MT1F;MT1G;MT1H |
|  | GO_bp:go_response_to_zinc_ion | 914.93 | 1.10E-16 | MT1E;MT1M;MT1A;MT1B;MT1F;MT1G;MT1H |
|  | GO_bp:go_cellular_response_to_cadmium_ion | 1787.03 | 3.40E-14 | MT1E;MT1A;MT1F;MT1G;MT1H |
|  | GO_bp:go_response_to_transition_metal_nanoparticle | 319.21 | 1.40E-13 | MT1E;MT1M;MT1A;MT1B;MT1F;MT1G;MT1H |
|  | GO_bp:go_cellular_response_to_inorganic_substance | 299.92 | 2.20E-13 | MT1E;MT1M;MT1A;MT1B;MT1F;MT1G;MT1H |
|  | GO_bp:go_negative_regulation_of_growth | 194.75 | 4.20E-12 | MT1E;MT1M;MT1A;MT1B;MT1F;MT1G;MT1H |
|  | GO_bp:go_response_to_cadmium_ion | 534 | 7.40E-12 | MT1E;MT1A;MT1F;MT1G;MT1H |
|  | GO_bp:go_response_to_metal_ion | 136.19 | 4.60E-11 | MT1E;MT1M;MT1A;MT1B;MT1F;MT1G;MT1H |
|  | GO_bp:go_response_to_inorganic_substance | 93.97 | 5.70E-10 | MT1E;MT1M;MT1A;MT1B;MT1F;MT1G;MT1H |
|  | GO_bp:go_regulation_of_growth | 70.4 | 4.00E-09 | MT1E;MT1M;MT1A;MT1B;MT1F;MT1G;MT1H |
|  | GO_cc:go_perinuclear_region_of_cytoplasm | 68.67 | 4.80E-09 | MT1E;MT1M;MT1A;MT1B;MT1F;MT1G;MT1H |
|  | GO_mf:go_zinc_ion_binding | 37.92 | 2.30E-07 | MT1E;MT1M;MT1A;MT1B;MT1F;MT1G;MT1H |
|  | GO_mf:go_transition_metal_ion_binding | 30.85 | 8.60E-07 | MT1E;MT1M;MT1A;MT1B;MT1F;MT1G;MT1H |
| Pallidum | GO_mf:go_triglyceride_lipase_activity | 584.38 | 6.80E-08 | PNLIPRP3;PNLIP;PNLIPRP1 |
| Putamen | GO_mf:go_triglyceride_lipase_activity | 584.38 | 6.80E-08 | PNLIPRP3;PNLIP;PNLIPRP1 |
| Pars triangularis | GO_bp:go_hyaluronan_metabolic_process | 408.94 | 2.10E-07 | ITIH1;ITIH3;ITIH4 |

*bp = biological process, cc = cellular component, mf = molecular function*

The most significant gene set was ‘*cellular response to zinc ion*’ for the precentral gyrus (OR = 4670.17; *p* = 3.9e-21). Here, the related genes all encode for metallothionein, which affect the cellular communication and signal transduction by binding to metal ions, such as zinc. All other gene sets detected for the precentral gyrus were also related to cellular response (in particular to metal ions) or regulation of growth. These results may suggest that the genetic liability in depression could be partially attributed to altered cell signalling in the precentral gurus resulting in detectable morphological changes in this region (though experimental validation will be required in order to confirm this relationship).

‘Triglyceride lipase activity’ was significant for both the pallidum and putamen, two regions that are part of the mesolimbic system which is associated with motivation and reward sensitivity. Relatedly, triglycerides are known to be metabolised within the mesocorticolimbic system and directly modulate sensitivity to rewards^33,34^, and it is possible that altered triglyceride activity within these regions might affect depression via an influence on reward processing.

Finally, ‘hyaluronan metabolic process’ was detected for the ‘pars triangularis’. Although this process has not been linked with depression in the past, hyaluronan is a major component of the extracellular matrix and plays an essential role in multiple cellular functions, including proliferation, migration, and neuronal plasticity, all of which are essential for normal brain functioning^35,36^.

While these analyses highlight gene sets that could potentially account for the genetic overlap between depression and these brain regions, a note of caution is necessary when interpreting these results. Given that we cannot correct for the correlation between genes, it is possible that the signal from proximally located genes could be confounded by LD, which could influence the enrichment in the event that those genes are also involved in the same gene sets. Genuine signal from proximally located genes does occur in reality however, particularly since genes with similar function may often be proximal to each other^37^, but without conditional analyses it is not possible to determine whether these results represent genuine findings or confounding. Nonetheless, these findings may still serve to generate actionable hypotheses about depression aetiology and prioritise gene sets that could be relevant for functional follow-up, specifically with a focus on the volumetric differences in these brain regions in relation to depression.

### Convergent evidence across molecular and neuroimaging modalities highlight *FLOT1*

We specifically looked for genes within which we found significant associations between depression and multiple endophenotypes as this could suggest potentially wide-ranging effects, making a gene particularly interesting for functional follow-up. From all analyses, the gene with the greatest number of significant associations was *FLOT1*, which was also the only gene where local genetic correlations were found on more than one level of function: the molecular level (*FLOT1* mRNA expression and splicing) and the neuroimaging level (left thalamus volume).

Alternative splicing for *FLOT1* mRNA transcripts were significantly genetically correlated with depression in 11 of the 18 analysed tissues (**Table 2**), with the most significant *FLOT1* association of all found with a splice variant in the spinal cord (which also represented the most significant association across all endophenotype analyses in this study). Though the strongest genetic correlation estimate within the *FLOT1* region was between depression and the volume of the left thalamus, for which the 95% confidence intervals for the *R*^2^ also included 1, which is consistent with the notion that the genetic signal in this locus is entirely shared between MDD and thalamus volume (**Table 2**).

**Table 2.** Significant associations detected within the FLOT1 cis-region (chr6:29,727,709-31,727,709)

| Level | Region / tissue | Splice variant | Rho | $R^2$ | $R^2_{lower}$ | $R^2_{upper}$ | P |
| --- | --- | --- | --- | --- | --- | --- | --- |
| Splicing | Brain spinal cord | 30740798:30741190 | **0.74** | 0.54 | 0.36 | 0.78 | 1.77e-16 |
|  | Brain cerebellar | 30740798:30741190 | **0.61** | 0.38 | 0.22 | 0.59 | 8.23e-13 |
|  | Brain caudate | 30741333:30741614 | **0.57** | 0.32 | 0.18 | 0.51 | 9.40e-13 |
|  | Brain cerebellar | 30740798:30741614 | **-0.56** | 0.32 | 0.18 | 0.50 | 2.16e-12 |
|  | Brain amygdala | 30740798:30741190 | **0.75** | 0.56 | 0.34 | 0.84 | 2.47e-12 |
|  | Brain frontal cortex b9 | 30741333:30741614 | **0.52** | 0.27 | 0.14 | 0.46 | 7.34e-10 |
|  | Brain hypothalamus | 30740798:30741614 | **-0.55** | 0.30 | 0.15 | 0.51 | 8.42e-10 |
|  | Thyroid | 30741333:30741614 | **0.53** | 0.28 | 0.13 | 0.50 | 2.87e-08 |
|  | Cells lymphocytes | 30740798:30741614 | **-0.50** | 0.25 | 0.11 | 0.46 | 1.04e-07 |
|  | Brain cerebellum | 30741867:30742147 | **-0.42** | 0.18 | 0.07 | 0.35 | 5.41e-07 |
|  | Brain nucleus accumbens | 30740798:30741190 | **0.48** | 0.23 | 0.09 | 0.43 | 6.40e-07 |
| Expression | Thyroid | - | **-0.45** | 0.20 | 0.10 | 0.33 | 1.23e-10 |
|  | Pituitary | - | **-0.68** | 0.46 | 0.23 | 0.77 | 2.59e-09 |
| Volume | Thalamus (left) | - | **0.84** | 0.70 | 0.30 | 1.00 | 3.56e-07 |

As expression QTL summary statistics for the thalamus is not present in the GTEx data, we also used GAMBA^38^ to examine *FLOT1* expression patterns based on data from the Allen Human Brain Atlas^39^. Here, we found the thalamus is among the top ten brain regions with the highest level of *FLOT1* expression (see **Suppl. Fig. 2** and **Suppl. Table 4**), though we note that no brain region showed a significantly higher expression level than expected by chance (*p* > .05).

Finally, we used StringDB^40^ to examine known and predicted interactions between FLOT1 and other proteins, which highlighted several transcripts of genes related to neuronal function, such as NGB (neuroglobin: involved in oxygen transport in the brain), APP (amyloid-beta A4 protein: triggers neuronal cell degeneration), and SLC6A3 (solute carrier family 6 member 3: sodium dependent dopamine transporter; see **Suppl. Fig. 3** for more detail).

## DISCUSSION

Depression is a heritable psychiatric disorder which represents one of the most significant disease burdens worldwide^3^. An improved understanding of its genetic component is integral to forming a clearer picture of its aetiology and developing effective treatments. But while extensive large-scale genetic association studies of depression have been performed, inferring function from GWAS results has been challenging, and so far, much of the biological mechanisms reflected in the genetic association signal remain elusive.

In this study, we have generated hypotheses on putative function of the depression GWAS signal in a novel manner, using local genetic correlation between depression and a range of molecular and brain imaging endophenotypes across different levels of function, with the purpose of elucidating relevant neurobiological paths through which depression variants could act. Considering the 1Mb cis region of genes as the loci of interest, we also perform endophenotype stratified gene-set enrichment analyses of the top 1% of genetically correlated genes, which provides additional context to any detected genetic correlations.

Using tissue specific gene expression and splicing endophenotypes, we show that the MDD GWAS signal peak located in the MHC is correlated with the genetically regulated expression and splicing of several genes, with *FLOT1* representing the gene with the greatest number of significant associations of all (detected in a total of 11 tissues: brain amygdala, brain caudate, brain cerebellar hemisphere, brain cerebellum, brain frontal cortex BA9, brain hypothalamus, brain nucleus accumbens, brain spinal cord, cells lymphocytes, pituitary, thyroid).

Analyses with regional brain volume endophenotypes identified significant local genetic correlations between MDD and regions primarily within and around the basal ganglia (basal forebrain, brain stem, caudate, lateral ventricle, pallidum, putamen, thalamus, ventral diencephalon), but also with frontotemporal regions (lingual gyrus, precentral gyrus, pars opercularis, transverse temporal gyrus) and two cerebellar vermal lobules (1/5 & 8/10). The emphasis on the basal ganglia differs from the functional follow-up analyses from Howard et al.^7^ (the summary statistics analysed here represent a large subset from their study). Using cell type specific enrichment from the gene analysis results, they implicated only the cortex, anterior cingulate cortex, and the frontal cortex. Given the central role of the basal ganglia in previous depression literature, this suggests that integration of multiple sources of data may aid the discovery potentially relevant targets which may be missed using traditional enrichment analysis. Though we emphasise that validation of our findings will be required in order to confirm a shared genetic aetiology between MDD and volumetric aberrations within these regions.

Further gene-based local genetic correlation analyses with the connectivity of seven brain networks identified four significant genetic correlations with the structural connectivity within the visual cortex within genes related to for example stress hormone regulation and circadian rhythm. Although higher order cognitive and emotional processing networks tend to be implicated in depression (such as the default mode, central executive, and ventral attention network), we note that power for all connectivity endophenotypes was generally low, and that these results are a likely consequence of the greater power of the visual network compared to the other networks.

In order to evaluate whether pleiotropic effects for any given endophenotype tended to be overrepresented within any gene sets, we performed stratified gene-set enrichment analyses of the top 1% of genes from the genetic correlation analyses for all analysed endophenotypes separately. Results from these analyses highlighted gene sets related to cellular response to metal ions and growth regulation for the precentral gyrus, triglyceride lipase activity for the palladium and putamen, as well as hyaluronan metabolism for the pars triangularis. While experimental validation will be necessary to confirm these findings, stratification by genetic correlation with relevant endophenotypes offers a powerful approach to contextualising discovered gene set associations, providing a more detailed picture of its potential relevance in the aetiology of the disorder.

Finally, of special interest in this study was to look for genes within which we observed genetic relations between depression and several endophenotypes, particularly across multiple levels of function, as it strengthens the evidence for the involvement of any particular gene and may also implicate such genes as having wide-ranging effects, making them particularly interesting for functional follow-up or pharmaceutical targeting. Here, the *FLOT1* gene stood out as the gene within which most significant associations were detected among all local genetic correlation analyses performed, and it was also the only gene where associations were found across two different modalities: molecular and neuroimaging (i.e. the *FLOT1* expression and splicing in multiple tissues, as well as the volume of the left thalamus).

*FLOT1* encodes a protein which is an integral membrane component of the caveola, and is likely a scaffolding protein involved in signal transduction^41^, vesicle trafficking^42^, and cell-cell adhesion^43^. Relatedly, it interacts directly with dopamine^44^ and serotonin transporters^45^ whose targets have been frequently implicated in depression^46–48^. Although *FLOT1* has itself received little attention in depression aetiology (unlike the monoamines which it regulates), it was recently flagged in a transcriptome wide-association study (TWAS) of depression that used an independent Chinese sample in addition to the European GTEx sample analysed in this study^10^. One recent experimental mouse study also examined the influence of down-regulated *FLOT1* expression in the hippocampus. There, decreased *FLOT1* was shown to exacerbate depressive behaviours in response to chronic corticosterone exposure^45^, which provides a suggested mechanism through which *FLOT1* variants might act in the context of MDD.

While little is currently known about *FLOT1* druggability, FLOT1 protein expression has been found to be responsive to exposure to valproic acid^49^, a drug which is frequently used to treat mood disorders (in particular bipolar disorder^50^). Efforts have also been made to target *FLOT1* pharmaceutically for the treatment of Alzheimer’s disease^51^, and since our results suggest that drugs targeting *FLOT1* may be promising to explore further in depression treatment, it may be worthwhile to examine whether such drugs could be efficacious for MDD as well.

A number of limitations should be kept in mind while interpreting the findings of this study. Firstly, the reliance on observational GWAS data greatly impacts any claims about causality or direction of effect that can be made based on the results of this study alone. While one may intuitively interpret genetic correlations between a lower and higher order phenotype in the direction of complexity, reverse causality or mediation by unobserved confounders are always possible. Similarly, given the strong LD between genes, the genetic signal for different genes are often correlated, meaning that significant associations from multiple proximal genes are expected even in scenarios when the true genetic signal arises from a single gene. This is a common problem when dealing with genetic data, and though conditional models could help prioritise likely causal genes, these were not available at the moment. Furthermore, it is also not possible to conclude that the endophenotypes implicated are exclusively correlated with depression, given that depression shows genetic overlap with multiple other neuropsychiatric disorders and traits^7,52,53^. An additional consideration is that since Bonferroni correction was used to correct for multiple testing, the significance threshold employed should be somewhat conservative since the number of independent associations will in practice be fewer than the total number of analyses (both when considering the correlation between genes, but also the endophenotypes). The reliance on summary statistics of European samples (which was done to maximise sample size), also means that the generalisability of our findings may not extend to populations of non-European ancestry.

The final, but perhaps most important limitation concerns the heterogeneity of MDD. That is, while MDD was analysed as a single, unified construct, in reality it is heterogeneous and syndromic, likely consisting of multiple sub-groups which differ with respect to symptom presentation and aetiology^54^. Since this study relied on publicly available summary statistics, there was no opportunity to account for this in the current analyses. But though our findings do not reflect the heterogeneous nature of MDD, they may nonetheless be relevant for MDD as a whole as they could point towards underlying biology that could be targetable in at least a notable proportion of MDD cases.

In conclusion, this study has demonstrated a novel way in which local genetic correlation may be applied to attempt to elucidate the function of the genetic signal in depression, using publicly available GWAS summary statistics from a range of relevant endophenotypes. Although the results presented cannot be interpreted as definite proof of any potential implied mechanism or causal path, they have generated several concrete and testable hypotheses about the possible ways in which genetic variants could influence depression, providing an important contribution to the body of research that aims to uncover its complex aetiology.

## METHODS

### Ethics statement

This study relied entirely on secondary analysis of publicly available summary statistics, for which ethical approval has been obtained by the primary researchers.

### LAVA overview

The LAVA method has been described in detail elsewhere^14^, but a brief overview is provided here. For a given locus, denote the standardized SNP genotype matrix for the locus as *X*, with *X* containing only SNPs for which summary statistics are available for all phenotypes to be analysed together. The genotype matrix is then projected onto its standardized principal components *W*, selecting the first *K* principal components, with *K* the smallest number such that *W* captured at least 99% of the variance in *X*. In a modification to the original LAVA implementation, *K* was subsequently constrained to be no greater than 80% of the smallest sample size of the phenotypes being analysed, which was necessary to maintain the stability of the parameter estimates when analysing the molecular data.

For each phenotype *p*, with phenotype variable *Y*_*p*_, LAVA then fits the multiple linear regression model *Y*_*p*_ = *Wδ*_*p*_ + *ε*_*p*_ if the phenotype is continuous (with *Y*_*p*_ standardized), or the equivalent multiple logistic regression model if the phenotype is binary. From this, the sampling distributions for the effect estimates 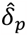 are obtained, which serve as the basis for the remainder of the analysis. In practice, the parameters of these sampling distributions are estimated using GWAS summary statistics for each of the phenotypes, together with genotype reference data to estimate LD. For each principal component *j*, the vector 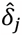 of its estimated effects across phenotypes is assumed to be distributed as 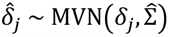. The matrix 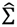 is the sampling covariance matrix, with diagonal elements equal to the sampling variance estimates 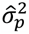 obtained from the univariate regression for each phenotype *p*. The off-diagonal elements reflect sampling covariance induced by possible sample overlap, which are computed using the intercepts from the corresponding bivariate LD score regression models^55^.

Defining the local genetic component matrix *G*, with columns *G*_*p*_ = *Wδ*_*p*_ for each phenotype, the aim of LAVA is to estimate and test the local genetic covariance matrix Ω = cov(*G*), with elements 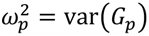 for each phenotype *p*, and *ω*_*pq*_ = cov(*G*_*p*_, *G*_*q*_) for each pair *p* and *q*. A method of moments estimator is used to obtain the corresponding estimate 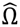. Univariate tests are performed for each phenotype to evaluate the presence of genetic signal for each phenotype in the locus, testing 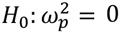.

Bivariate tests of local genetic correlation between *p* and *q* are performed to evaluate genetic overlap in the locus, testing *H*_0_ : *ω*_*pq*_ = 0. The local genetic correlations are defined as 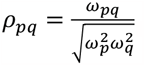, with the estimate 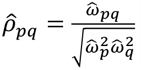. Multivariate models for more than two phenotypes can similarly be evaluated, using transformations of 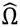 to obtain multiple linear regression and partial correlation models for *G*.

### Meta-analysis of local genetic components

The LAVA model was extended to implement a form of meta-analysis at the level of genetic components, allowing for the estimation and testing of genetic correlations of a composite genetic component *G*_*M*_ with the genetic components of other phenotypes. This can be used to perform LAVA analysis for linear combinations of phenotypes, using only the summary statistics for those individual phenotypes. We present this model here for the special case where *G*_*M*_ is a composite of the genetic components of two phenotypes, *G*_1_ and *G*_2_, correlated with that of a single other phenotype, *G*_*D*_, but the framework generalized to arbitrary numbers of phenotypes.

The composite genetic component is specified as a weighted sum *G*_*M*_ = *w*_1_*G*_1_ + *w*_2_*G*_2_ = *w*_1_*Wδ*_1_ + *w*_2_*Wδ*_2_ = *Wδ*_*M*_, with weights *w*_1_ and *w*_2_ and with *δ*_*M*_ = *w*_1_*δ*_1_ + *w*_2_*δ*_2_. This shows that this approach is essentially equivalent to meta-analysing the genetic effects before running the LAVA analysis, provided the same weights are used. Note that the absolute scaling of the weights does not matter, only the scaling of *w*_1_ relative to *w*_2_, since changes in absolute scaling will cancel out when computing genetic correlations of *G*_*M*_ with any other phenotype.

The full local genetic covariance Ω for *G*_1_, *G*_2_ and *G*_*D*_ can be defined and estimated using the standard LAVA model as outlined above, and this can be used to define and estimate the transformed genetic covariance matrix 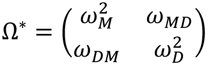. Since *G*_*M*_ is a linear combination of *G*_1_ and *G*_2_, the elements of Ω^∗^ have the form 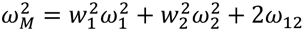 and *ω*_*DM*_ = *ω*_*MD*_ = *w*_1_*ω*_1*D*_ + *w*_2_*ω*_2*D*_. The estimate 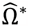 can therefore be computed directly from 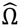, and the transformed sampling covariance matrix 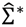 can be computed from the full 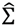 in the same way.

Subsequent analysis proceeds in the same way as in the core LAVA model. An inverse-variance weighting approach is used to set the weights, with 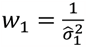 and 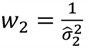. The meta-analysis procedure has been added to the LAVA R package and can be called via the ‘meta.analyse.locus()’ function (more detail can be found in the function manual).

### Summary statistics and genotype reference data

Summary statistics for depression were downloaded from the GWAS atlas (https://atlas.ctglab.nl)^56^, and represent the subset from the Howard et al. (2019)^7^ GWAS meta-analysis of depression excluding the 23andMe samples (*N* = 500,199). The cis-eQTL and cis-sQTL summary statistics were obtained from GTEx v8 (https://gtexportal.org/home/datasets), and the brain volume GWAS summary statistics by Zhao et al. (2021)^19^ were downloaded from https://www.med.unc.edu/bigs2/data/gwas-summary-statistics/. Finally, the network connectivity GWAS summary statistics by Tissink et al. (2023) are available at https://ctg.cncr.nl/software/summary_statistics/.

All GWAS summary statistics are based on participants of European genetic ancestry, and we used genotype reference data from for the European subset of the 1000 Genomes (phase 3)^57^ sample for the estimation of SNP LD for the LAVA analyses. We filtered all summary statistics to SNPs having a minor allele frequency of at least .5% in the 1000 Genomes data in order to exclude SNPs with a very low frequency.

### Local genetic correlation analysis with molecular phenotypes

In order to limit the number of performed tests downstream, and thus the multiple testing burden, we pre-selected GTEx tissues that we judged to be of greatest potential relevance to depression based on the existing literature, which has implicated primarily the brain^9,17^, but also the endocrine (adrenal gland, pituitary, thyroid) and immune systems (whole blood, EBV cells)^18^. As loci, we used the cis-region of all protein coding genes (*N* = 18,380), including all SNPs that were within one megabase of the gene transcription start site.

For every tissue and type (i.e. gene expression or splicing), LAVA univariate tests were performed for both depression and the molecular phenotype. For each gene for which sufficient genetic signal was present for both (univariate *p*-values < 1e-4), the local bivariate genetic correlation of the molecular phenotype with depression was estimated and tested. The sampling correlations in the bivariate analyses were all assumed to be negligible, and the off-diagonal elements of 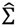 were therefore set to zero. Bonferroni correction was used to account for multiple testing, correcting for the total number of local genetic correlations with depression tested across all bivariate local *r*_g_ analyses in the paper (that is, including all molecular and brain imagining endophenotypes), resulting in a significance threshold of 0.05 / 72,701 = 6.88e-7.

### Local genetic correlation analysis with brain imaging phenotypes

For the LAVA analyses of the regional brain volume and network connectivity phenotypes, we again defined our loci as using all protein coding genes (*N* = 18,380), adding one megabase windows around the transcription start sites to obtain the entire cis-regions. As for the molecular phenotypes, for each brain phenotype univariate tests were performed for all genes, selecting genes for bivariate analysis if univariate *p*-values for both the brain phenotype and depression for that gene were lower than 1e-4.

For the regional brain volume phenotypes, an extra step was included to account for the potentially quite strong genetic correlations between the hemispheric homologous regions (i.e. regions for which there are corresponding but separate regions from both left and right hemispheres). For every homologous region and gene, if both the left and right hemisphere for that region were selected for bivariate analysis, the local genetic correlation *ρ*_*lr*_ between the two hemispheres was first computed for that region. If either 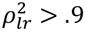, or the 95% confidence interval of 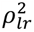 included 1, the genetic components of the two hemispheres were meta-analysed (see also *Meta-analysis of genetic components* above), and the local genetic correlation of this meta-analysed genetic component with that of depression was estimated and tested. Otherwise, local genetic correlations for that region with depression were analysed separately for the left and right hemisphere.

In the bivariate analyses, to account for sample overlap, LD score regression^55^ was used to obtain estimates of the sampling correlations between depression and the brain imaging phenotypes, as well as between the left and right hemisphere regional brain volumes for each region. These were provided to LAVA to set the off-diagonal elements of 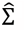. As with the molecular phenotypes, a Bonferroni-corrected significance threshold of 0.05 / 72,701 = 6.88e-7 was used, correcting for the total number of local genetic correlations tested across all genes and endophenotypes analysed in this study (i.e. both molecular and brain imaging).

### Endophenotype-stratified gene-set enrichment analysis

We examined whether the endophenotype-specific genes implicated through local *r*_g_ analysis with depression were enriched within any biological annotated pathways, by conducting endophenotype-stratified gene-set enrichment analyses. Thus, for each analysed endophenotype separately, the top 1% of genes from local *r*_g_ analyses with depression were subjected to a Fisher’s exact test in order to evaluate over-representation within 7,246 MSigDB v6.2^15^. Bonferroni correction was used to account for multiple testing, correcting for the number of enrichment analyses performed across all phenotypes, resulting in a significance threshold of 0.05 / 28,959 = 1.73e-6 (note that for each endophenotype, only gene sets which included at least one evaluated gene from the local *r*_g_ analyses were tested).

## Data Availability

All data produced in the study are available online at https://github.com/josefin-werme/lava-mdd-endo-2023

## Data availability

All analyses in this study relied on publicly available data from GWAS atlas https://atlas.ctglab.nl, GTEx https://gtexportal.org/home/datasets, brain volume GWAS https://www.med.unc.edu/bigs2/data/gwas-summary-statistics/, network connectivity GWAS https://ctg.cncr.nl/software/summary_statistics/, and MSigDB https://www.gsea-msigdb.org/gsea/msigdb/collections.jsp. The locus file used for all the LAVA analyses can be downloaded from https://github.com/josefin-werme/lava-mdd-endo-2023.

## Code availability

Analysis scripts to generate the results in this manuscript are publicly available at https://github.com/josefin-werme/lava-mdd-endo-2023. The LAVA software is implemented as an R package (https://github.com/josefin-werme/lava) and contains the relevant functions used for the local *r*_g_ analyses in this study.

## Acknowledgements

This work was funded by COSYN (Comorbidity and Synapse Biology in Clinically Overlapping Psychiatric Disorders: Horizon 2020 Program of the European Union under RIA grant agreement 667301, to D.P.), the Netherlands Organization for Scientific Research (NWO: VICI 435-14-005, to D.P.), NWO Gravitation: BRAINSCAPES: A Roadmap from Neurogenetics to Neurobiology (Grant No. 024.004.012, to D.P.), and a European Research Council advanced grant (Grant No, ERC-2018-AdG GWAS2FUNC 834057, to D.P.). The work of S.C.d.L. was supported by ZonMw, the Hague, The Netherlands, project 09120011910032 REMOVE and the European Research Council, Brussels, Belgium, Advanced Grant 101055383 OVERNIGHT. C.A.d.L. is funded by Hoffman-La Roche. The work of M.H. was supported by a VIDI (452-16-015) grant from the Netherlands Organization for Scientific Research (NWO) and an ERC Consolidator of the European Research Council (101001062). Analyses were carried out on the Genetic Cluster Computer hosted by the Dutch National computing and Networking Services SURFsara and financed by the Netherlands Organization for Scientific Research (NWO: 480-05-003), the VU University (Amsterdam, The Netherlands) and the Dutch Brain Foundation.

## Author contributions

J.W., C.A.d.L. & D.P. conceived of the study. J.W. performed the analyses and drafted the manuscript with input from E.T. and C.A.d.L. All authors contributed to revision of the manuscript and interpretation of results.

