## Supplementary Figures for "Local genetic correlation analysis links depression with molecular and brain imaging endophenotypes"

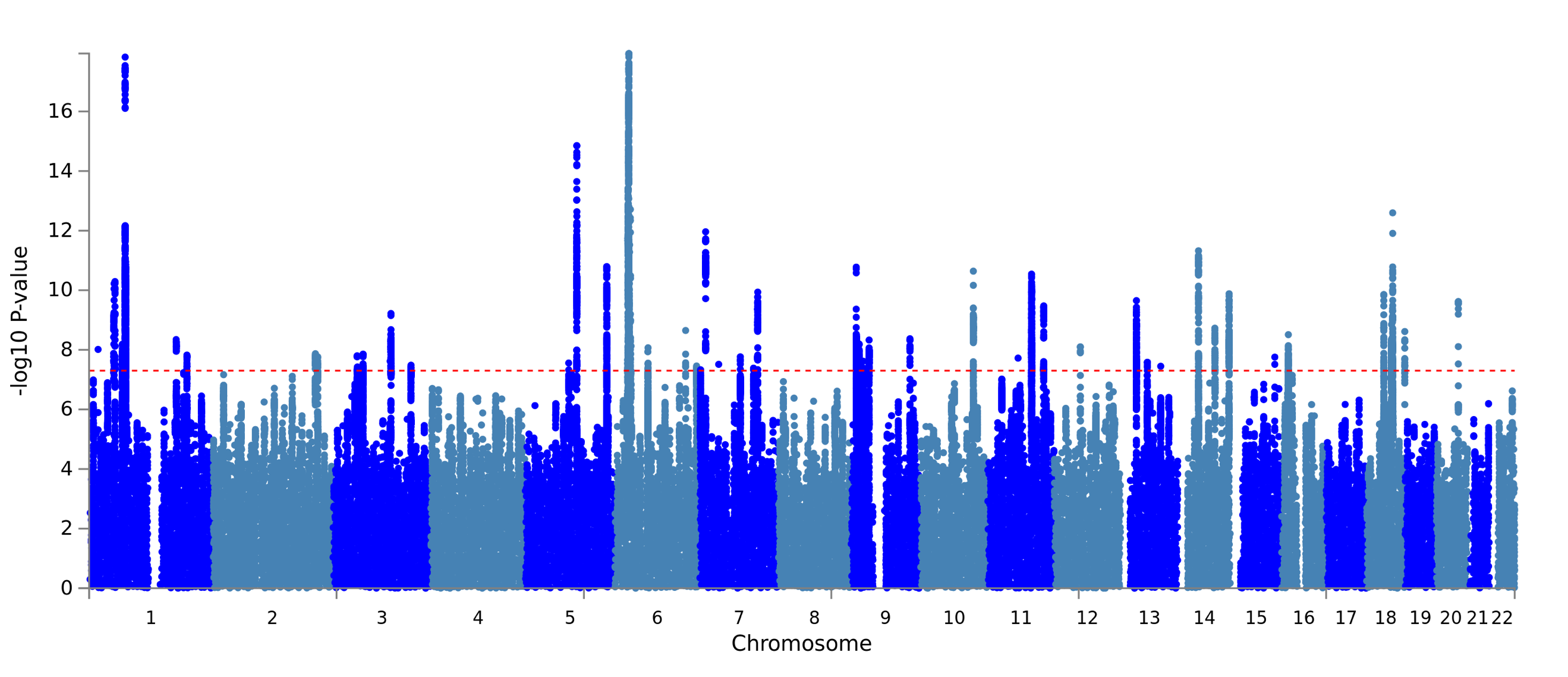

***Supplementary Figure 1 Manhattan plot for the depression summary statistics.*** *Downloaded from the GWAS atlas*^1^ *at https://atlas.ctglab.nl/traitDB/4293. Represents the depression GWAS summary statistics from Howard et al.*^2^ *(representing the European subset of N=* *500,199) which were analysed in this study.*

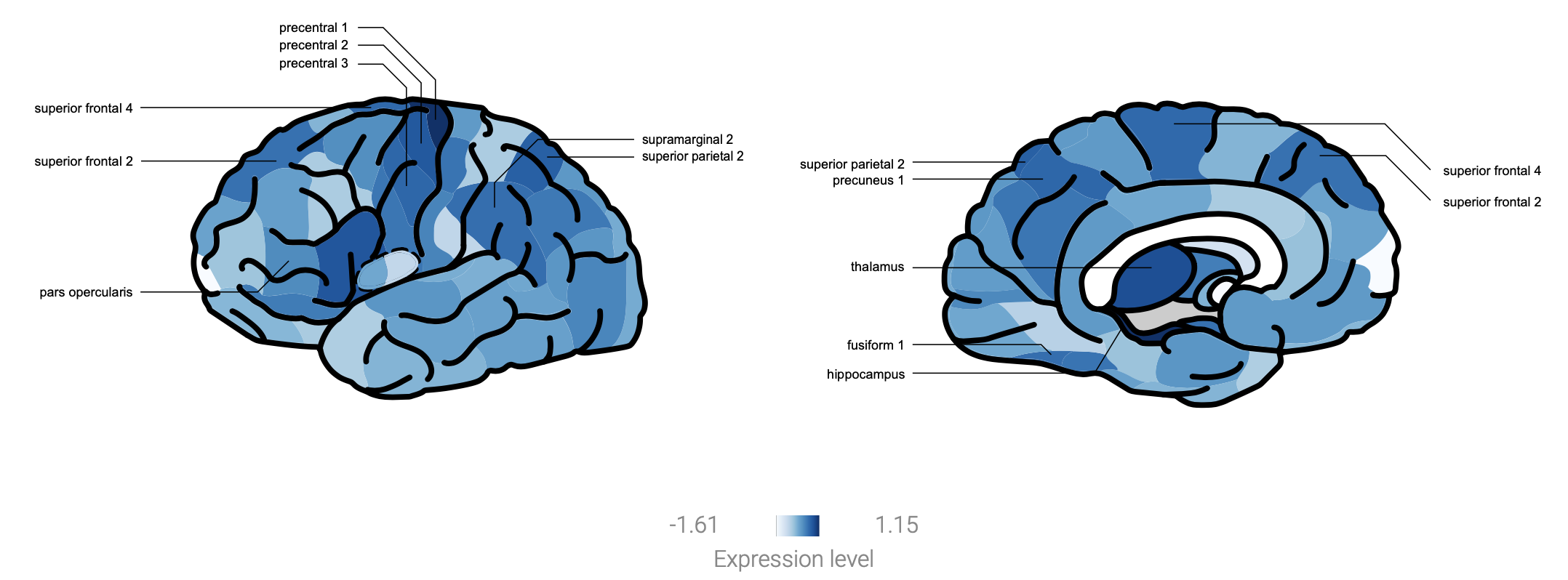

***Supplementary Figure 2 Illustration of FLOT1 gene expression levels from the Allen Human Brain Atlas (generated with GAMBA***^3^***.*** *The top 10 regions showing the highest levels of expression for the FLOT1 gene are labelled (see* ***Suppl. Table 4*** *for the expression levels per region), though note that no brain region showed significantly higher FLOT1 expression than expected by chance (p < .05).*

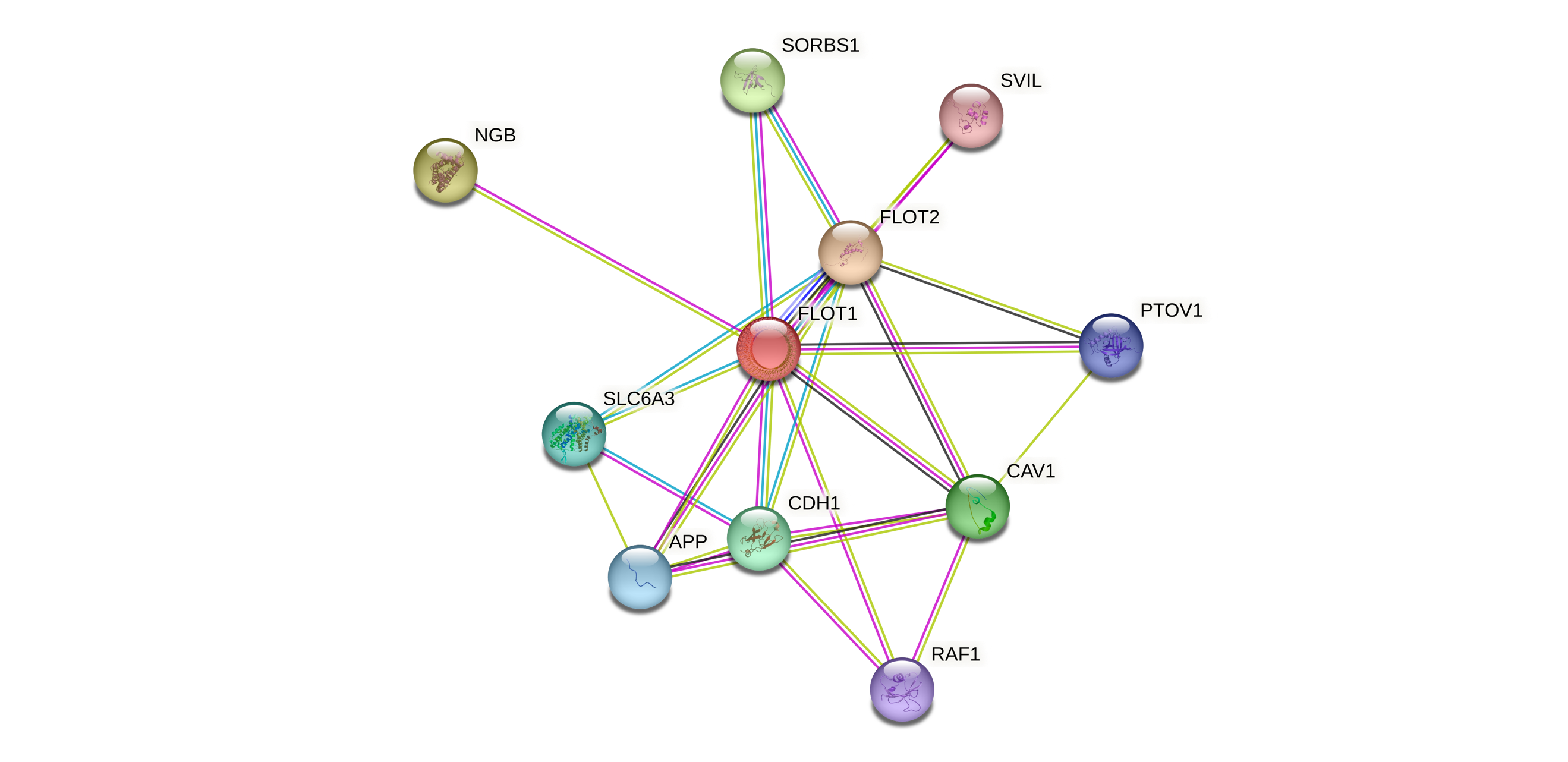

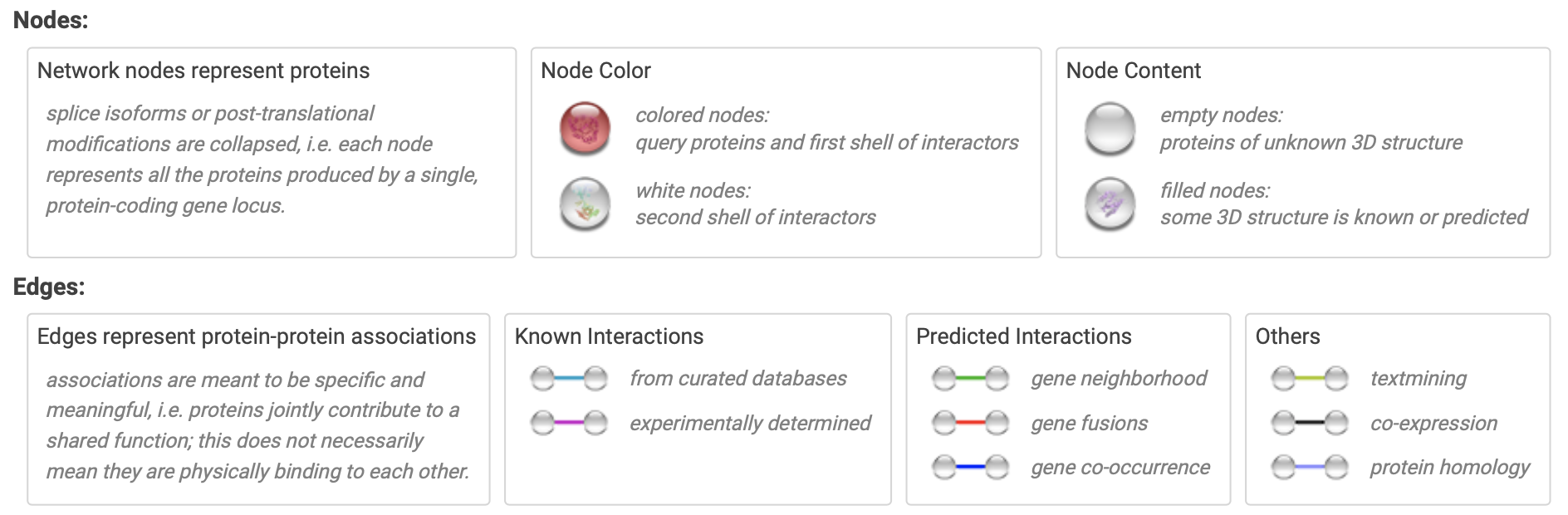

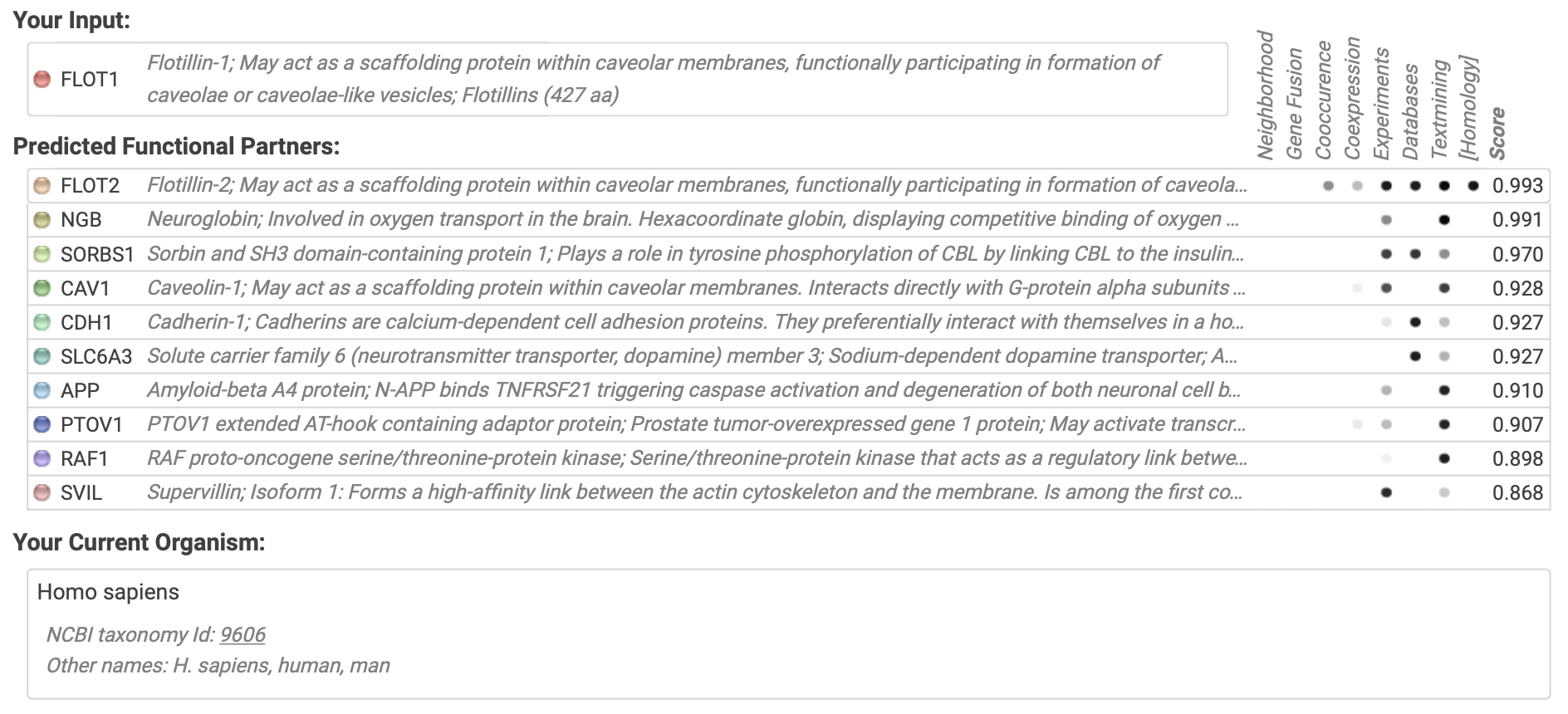

***Supplementary Figure 3 StringDB protein-protein interaction network for FLOT1.*** *Generated via https://string-db.org^4^.*
